# A comprehensive accuracy assessment of Samsung smartwatch heart rate and heart rate variability

**DOI:** 10.1101/2022.04.29.22274461

**Authors:** Fatemeh Sarhaddi, Kianoosh Kazemi, Iman Azimi, Rui Cao, Hannakaisa Niela-Vilén, Anna Axelin, Pasi Liljeberg, Amir M. Rahmani

## Abstract

**Background:** Photoplethysmography (PPG) is a low-cost and easy-to-implement method to measure vital signs, including heart rate (HR) and heart rate variability (HRV). The method is widely used in various wearable devices. For example, Samsung smartwatches are PPG-based open-source wristbands used in remote well-being monitoring and fitness applications. However, PPG is highly susceptible to motion artifacts and environmental noise. A validation study is required to investigate the accuracy of PPG-based wearable devices in free-living conditions.

**Objective:** We evaluate the accuracy of PPG signals – collected by the Samsung Gear Sport smartwatch in free-living conditions – in terms of HR and time-domain and frequency-domain HRV parameters against a medical-grade chest electrocardiogram (ECG) monitor.

**Methods:** We conducted 24-hours monitoring using a Samsung Gear Sport smartwatch and a Shimmer3 ECG device. The monitoring included 28 participants (14 male and 14 female), where they engaged in their daily routines. We evaluated HR and HRV parameters during the sleep and awake time. The parameters extracted from the smartwatch were compared against the ECG reference. For the comparison, we employed the Pearson correlation coefficient, Bland-Altman plot, and linear regression methods.

**Results:** We found a significantly high positive correlation between the smartwatch’s and Shimmer ECG’s HR, time-domain HRV, LF, and HF and a significant moderate positive correlation between the smartwatch’s and shimmer ECG’s LF/HF during sleep time. The mean biases of HR, time-domain HRV, and LF/HF were low, while the biases of LF and HF were moderate during sleep. The regression analysis showed low error variances of HR, AVNN, and pNN50, moderate error variances of SDNN, RMSSD, LF, and HF, and high error variances of LF/HF during sleep. During the awake time, there was a significantly high positive correlation of AVNN and a moderate positive correlation of HR, while the other parameters indicated significantly low positive correlations. RMSSD and SDNN showed low mean biases, and the other parameters had moderate mean biases. In addition, AVNN had moderate error variance while the other parameters indicated high error variances.

**Conclusion:** The Samsung smartwatch provides acceptable HR, time-domain HRV, LF, and HF parameters during sleep time. In contrast, during the awake time, AVNN and HR show satisfactory accuracy, and the other HRV parameters have high errors.

## Introduction

Heart rate (HR) and heart rate variability (HRV) are physiological parameters reflecting autonomous nervous system regulations and general well-being. HR shows the number of heartbeats per minute, and HRV indicates the variation of time between two consecutive heartbeats or interbeat intervals (IBIs) [1]. Various HRV parameters can be extracted from IBIs, such as average normal IBIs (AVNN), standard deviation of normal IBIs (SDNN), and root mean square of the successive difference (RMSSD). HR and HRV parameters can provide insight into cardiovascular and autonomic nerve dysfunction [2]. Studies in the literature show the relationship between HRV parameters and different health issues such as diabetes [3], hypertension [4], depression [5], and autonomic imbalance [6]. Moreover, HRV parameters are associated with mental and physiological stress [7, 8], and sleep quality [9].

HR and HRV can be monitored using noninvasive methods such as Electrocardiography (ECG) and Photoplethysmography (PPG). ECG is the golden standard for HR and HRV parameters monitoring used in clinical trials. The method measures the electrical activity of the cardiovascular system using electrodes connected to the skins. However, it cannot be employed in home-based and/or long-term monitoring when people are engaged in different activities. Alternatively, PPG is another non-invasive method for HR and HRV monitoring. PPG is an optical method that utilizes a light emitter and a photodetector to measure the volumetric variations of blood flow [10]. PPG is a low-cost and convenient method implemented in many clinical and commercial wearable devices [11–13].

Recently, several PPG-based wearable devices have been proposed for health parameters monitoring in everyday life settings. Several studies leveraged different wearable devices, such as Samsung Gear Sport, Apple Watch, Fitbit, and Garmin Vivosmart, for health monitoring in different population-based groups [12–14]. With advancements in technology, it is expected that the use of such wearable devices will grow further as they become smaller and lighter with longer battery life. However, PPG-based wearable devices are prone to environmental noises and motion artifacts (when users engage in various physical activities). These noises are inevitable in everyday life settings and affect the signal quality, resulting in poor/invalid health parameters extraction [15]. Therefore, using commercial PPG-based wearable devices for HR and HRV monitoring necessitates accuracy assessment, especially if the devices are used for health monitoring applications.

Several studies investigated the validation of HR measurements using wristbands in various situations across different population groups. In [16], the authors evaluated the HR of several wristbands – including Apple Watch, Basis Peak, Fitbit Surge, Microsoft Band, Mio Alpha 2, PulseOn, and Samsung Gear S2 – during different physical activities. Other studies validated the HR extracted from Garmin Forerunner [17], the everlast smartwatch [18], Fitbit Charge HR [19], Empatica E4 [20], and Basis peak [21]. These studies showed the high accuracy of PPG-based wristbands for HR monitoring during resting and low-intensity activity in laboratory settings. The results also showed a decrease in the accuracy of HR when the intensity of activity increased. However, these studies are restricted to certain physical activities in laboratory settings. They are also limited to HR measurements. The accuracy of HR and HRV parameters is affected by different factors. For example, the accuracy of RMSSD can be affected by a distortion in a small part of the signals. However, SDNN accuracy can be impacted by outliers affecting the IBIs variations [22]. These characteristics of HRV parameters indicate the need to validate the accuracy of HRV parameters individually.

Studies evaluated the accuracy of HRV parameters extracted from wristbands and smartwatches including Apple Watch [23], Empatica E4 [24, 25], Microsoft band 2 [26], and the Wavelet wristband [27] against medical-grade ECG device. These studies indicated high accuracy of the smartwatches and wristbands in terms of HR and HRV parameters while the participants were resting. They also showed that motion artifacts highly affect the reliability of HRV parameters. However, these studies were limited to short-term data collection –less than one hour– in laboratory settings [20, 25, 26]. In addition, the majority of the previous works collected data only in seated positions [23, 24, 27].

We believe that there is a need to evaluate the accuracy of the smartwatch in everyday life settings where participants can engage in different activities and conditions. Such evaluation should also comprehensively assess the accuracy of time-domain and frequency-domain HRV parameters extracted from the raw PPG signals.

In this paper, we assess the validity of the Samsung Gear Sport smartwatch in terms of HR and several HRV parameters. The evaluation is performed against a medical-grade chest ECG monitor in a 24-hours continuous free-live setting monitoring. The data from 28 individuals are included in the evaluation. We use PPG and ECG signals collected from the Samsung smartwatch and ECG monitor to extract HR, AVNN, RMSSD, SDNN, pNN50, LF, HF, and LF/HF ratio in five-minute windows. We evaluate the parameters during sleep time and awake time. The evaluation is performed using a linear regression method, the Pearson correlation coefficient, and the Bland–Altman plot. Finally, We discuss the validity of the parameters based on the obtained results in everyday settings. In summary, the main contributions of this paper are as follows:

1. We investigate the validity of PPG signals acquired by the Samsung Gear Sport smartwatch in terms of HR and HRV parameters compared with a medical-grade chest ECG monitor
2. We conduct a 24-hours study in which 28 healthy participants are monitored remotely and continuously.
3. We analyze the HR and HRV parameters in five-minute windows during sleep-time and awake-time using the linear regression method, the Pearson correlation coefficient, and the Bland–Altman plot.

## Method

### Study design

An observational study was conducted to assess the validity of HR and HRV parameters collected under free-living conditions via Samsung Gear Sport smartwatches. The assessment was performed in comparison with an ECG monitor as the golden standard. The study included a convenience sample of healthy individuals recruited in Southwest Finland in July-August 2019.

### Participants and recruitment

Forty-six healthy individuals between the age of 18 and 55 were recruited to participate in this study. The inclusion criteria were individuals who 1) were able to use wearable devices for 24 hours, 2) had no diagnosed cardiovascular disease, 3) had no symptoms of illness during the recruitment time, and 4) had no restrictions in physical activities.

In a face-to-face meeting with researchers, the eligible participants were informed about the purposes of the study, the procedure, and the instructions to use the devices. After the written informed consent, the devices – a Samsung Gear sport smartwatch [28] and a shimmer3 ECG device [29] – were delivered to the participants. The participants were asked to wear the wearable devices for 24 hours continuously while engaging in their daily routines and log their sleep and awake time.

After the data collection, we excluded the data of 18 participants due to technical and practical issues during the monitoring, for example, ECG electrodes were not adequately attached to the skin, and participants forgot to log their sleep and awake time. Therefore, the data of 28 participants (i.e., 14 female and 14 male) were included in the analysis. Table 1 summarizes the background information of the participants.

**Table 1.** Participants background information n=27 (one participants didn’t fill the background questionnaire

| Parameters | Values |
| --- | --- |
| Age, years, mean (SD) | 32.5 (6.6) |
| BMI, mean (SD) | 25.4 (5.2) |
| Exercise |  |
| Almost daily | 8 |
| A few times a week | 13 |
| Once a week or fewer | 6 |
| Education |  |
| Primary school | 1 |
| High school | 6 |
| College | 6 |
| University | 14 |
| Employment status |  |
| Working | 22 |
| Student | 1 |
| Unemployed | 3 |
| Other | 1 |

### Research ethics

The study was conducted according to the ethical principles based on the Declaration of Helsinki and the Finnish Medical Research Act (No 488/1999). The study protocol received a favorable statement from the ethics committee (University of Turku, Ethics committee for Human Sciences, Statement no: 44/2019). The participants were informed about the study, both orally and in writing, before the written informed consent was obtained. Participation was voluntary, and all the participants had the right to withdraw from the study at any time and without giving any reason. To compensate for the time used for the study, each participant got a gift card to the grocery store (20 euro) at the end of the monitoring period when returning the devices.

### Data collection

The data collection included two wearable devices and self-report and background questionnaires. The participants were asked to wear a Samsung Gear Sport smartwatch on the wrist of their non-dominant hand. Moreover, they were asked to wear a Shimmer3 ECG device using a chest strap. The ECG was collected via four limb electrodes placed on the torso (i.e., left arm, right arm, left leg, and right leg). More details can be found in [30]. We also used self-report questionnaires by which the individuals logged their sleep and non-wear time. In our analysis, the self-report data were used to extract the sleep and awake time.

The Shimmer3 ECG device was selected to measure ECG as the gold standard method in our assessment. The Shimmer ECG is a compact and lightweight device that can be configured to measure ECG, accelerometer/gyroscope data, and skin temperature continuously [29]. The device also has sufficient internal memory and battery life for 24 hours of continuous data collection. We configured the Shimmer device to collect data with the sampling frequency of 512HZ, used in clinical trials to extract HR and HRV parameters accurately [31]. The data were stored on the device during the monitoring and were transferred to our cloud server after the monitoring for the analysis. In this study, Lead II ECG (right arm - left leg) was selected to extract the cardiac rhythm accurately.

The Samsung Gear Sport watch is a commercial open-source smartwatch that enables remote health monitoring [28]. The smartwatch provides PPG signals and gyroscope/accelerometer data at the sampling frequency of 20Hz. The watch runs an open-source Tizen operating system and has a built-in inertial measurement unit (IMU) to extract physical activity and sleep data. We developed a customized data collection application for the watch to collect 16 minutes of PPG signals every 30 minutes continuously. In the analysis, we removed the first minute of each PPG record, as it was unreliable due to sensor calibration. With this setup, the smartwatch’s battery life was more than 24 hours, so no battery charging was needed during the monitoring. The data was stored in the watch’s internal storage during the monitoring. Similar to the Shimmer device data, we transferred the watch’s data to the cloud after the monitoring.

### Data analysis

In this section, we describe HR and HRV extraction from the collected PPG and ECG signals. We used short-term HRV analysis, which considers five-minute windows of signals for the analysis [22, 32]. The short-term HRV analysis was selected based on the duration of PPG recordings (16 minutes per 30 minutes). We then outline the statistical analysis methods used in this study.

#### HR and HRV extraction pipeline

The HR and HRV extraction pipeline consists of four steps: filtering, peak detection, abnormal peak removal, and feature extraction (see Fig 1). The raw PPG and ECG signals are divided into five-minute segments. In the following, we provide a detailed description of each step:

**Fig 1.**
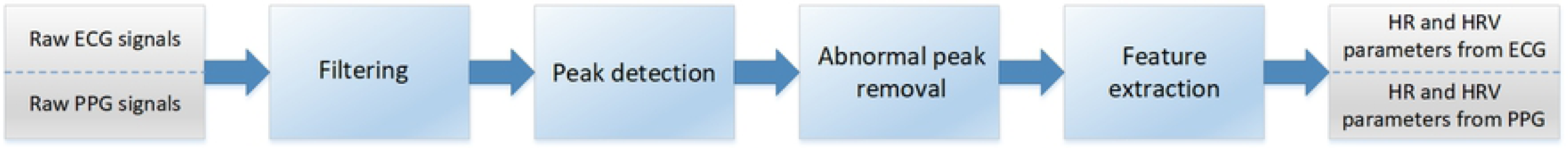
HR and HRV extraction pipeline

##### 1) Filtering

In this step, we remove frequencies that are out of the human heart rate range. We used a 5-order high-pass Butterworth filter with a cutoff frequency of 0.5 Hz for PPG signals and a bandpass Butterworth filter with 0.5 and 100 Hz cutoff frequencies for ECG signals. The cutoff frequencies were selected based on valid HR range and input signals’ frequencies.

##### 2) Peak detection

This step finds the peaks corresponding to the heartbeat in PPG and ECG signals.

###### PPG peak detection

We used a deep-learning-based method introduced by Kazemi *et al*. [33] for PPG peak detection. The method is enabled by a dilated Convolutional Neural Networks (CNN) architecture. The dilated convolutions provide a large receptive field, enhancing the efficiency of time-series processing with CNNs. The model outputs the probability of a signal point being a systolic peak. A peak finder function then detects the peaks’ locations in the signal. The peak finder function first makes a list of all points in the signals with a probability value higher than a pre-defined threshold (selected experimentally). Then, this function extracts the peaks’ locations using a local maximum finder.

###### ECG peak detection

We developed a two-round peak detection algorithm to locate peaks in ECG signals. In the first round, the algorithm computes the average value of filtered ECG signals in a 5-minute window. Then, it uses this average value as a threshold to detect all possible peaks, including the real and false peaks. In the second round, the algorithm computes the average value of all detected peaks from the first round as a new threshold. By using the new threshold value and the heartbeat range (i.e., 20-200 beats per minute), the undetected R peaks are added, and false peaks are removed. Our peak detection method obtains higher accuracy in comparison with Pan-Tompkins [34] and Hamilton [35] algorithms. Fig 2 shows a sample of the peak detection results in a 30 seconds segment of PPG and corresponding ECG signals.

**Fig 2.**
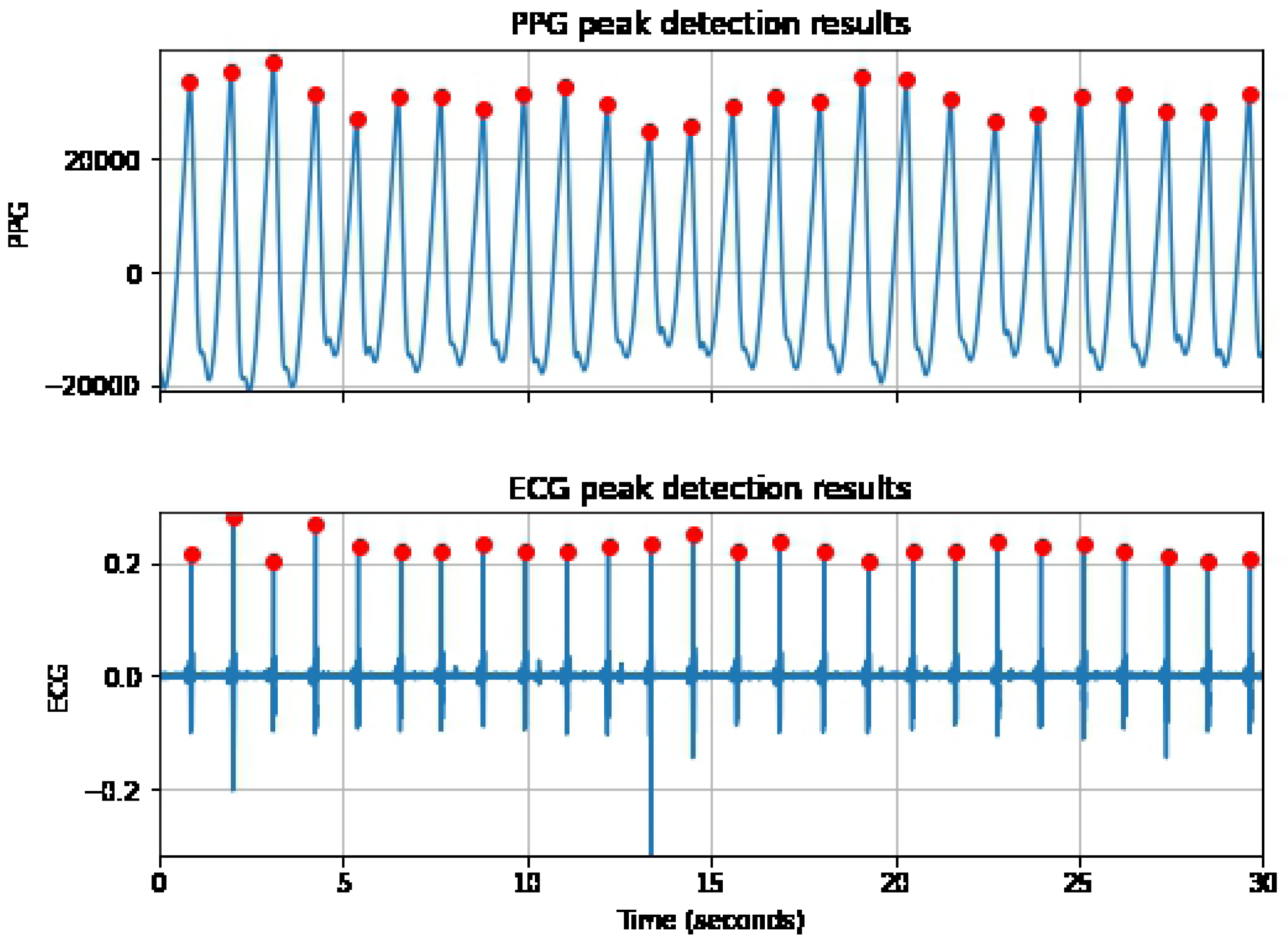
The peak detection results for a 30 seconds segment of PPG and ECG signals.

##### 3) Abnormal peak removal

We used a rule-based method to remove invalid peaks extracted in the previous step. The invalid peaks removal rules are as follows:

- We assume that the minimum and maximum heart rates are 20 and 200 beats per minute. Accordingly, the minimum and maximum peak-to-peak distances are 3000 and 300 milliseconds.
- If the variation in NN intervals exceeds 20% of the average NN intervals, the exceeding part is removed. Accordingly, if more than 50% of the total NN intervals were removed, then the result of the entire 5-minute segment is not considered.

##### 4) Feature extraction

In this step, we extracted HR and HRV parameters from normal interbeat intervals (NN intervals). The extracted time-domain HRV parameters are AVNN, SDNN, RMSSD, Percentage of successive NN intervals that differ by more than 50 ms (PNN50), and the frequency-domain parameters are low-frequency power (LF), high-frequency power (HF), and LF to HF ratio (LF/HF). Table 2 indicates the HRV parameters used in this study.

**Table 2.** Time domain and Frequency domain HRV features and the descriptions

| Feature | Units | Description |
| --- | --- | --- |
| Time domain features |  |  |
| AVNN | ms | Mean of NN intervals |
| SDNN | ms | Standard deviation of NN intervals |
| RMSSD | ms | Root mean square of successive NN interval differences |
| pnn50 | % | Percentage of successive NN intervals that differ by more than 50 ms |
| Frequency domain features |  |  |
| LF power | $s^2$ | Power in low frequency band (0.04 – 0.15) |
| HF power | $s^2$ | Power in high frequency band (0.15 – 0.4) |
| LF/HF | % | LF to HF ratio |

#### Statistical analysis

We investigated the linear relationship between the smartwatch and Shimmer3 by performing the Pearson Correlation coefficient test on extracted parameters from two devices. We also applied a linear regression analysis method to assess the accuracy of the smartwatch’s HR and HRV parameters. We used the Samsung Watch’s data points (HR and HRV parameters) to fit the linear regression line. Then, we computed the R-squared value (*r*^2^) using the regression line and corresponding ECG data points to evaluate the closeness of baseline data to the smartwatch’s fitted regression line.

Moreover, the Bland-Altman analysis was utilized to illustrate and estimate the agreement between the PPG and ECG results. The Bland-Altman analysis provides mean bias, standard deviation, and ±95% confidence intervals (CI) based on the differences between the Samsung Watch and Shimmer3. We leveraged python libraries including Scipy [36] and Statsmodels [37] to implement the statistical analysis.

## Results

We validated the PPG data collected via the Samsung smartwatch against the ECG data of the Shimmer device in free-living conditions. The analysis includes the data collected from 28 participants (i.e., 14 females and 14 males). We first assess the HR and HRV parameters derived from five-minute segments collected during sleep. Then, the five-minute PPG segments collected during the awake time are evaluated.

### Comparisons of HR and HRV parameters of Samsung smartwatch and Shimmer3 in 5-minute time windows during sleep time

The sleep duration was acquired from the self-report questionnaires collected during the monitoring. We obtained the correlation between the HR and HRV parameters of the smartwatch and Shimmer3 in 5-minute segments. Table 3 indicates the Pearson correlation coefficient with the corresponding P-values, 95% Confidence Interval, and mean biases of the HR and HRV parameters. As shown in Table 3, the HR, AVNN, SDNN, and PNN50 between the Samsung smartwatch and Shimmer3 are highly correlated. The correlation values of the RMSSD, LF, and HF are still high (positive) but slightly lower. The LF/HF ratio value shows a moderately positive relationship.

**Table 3.**
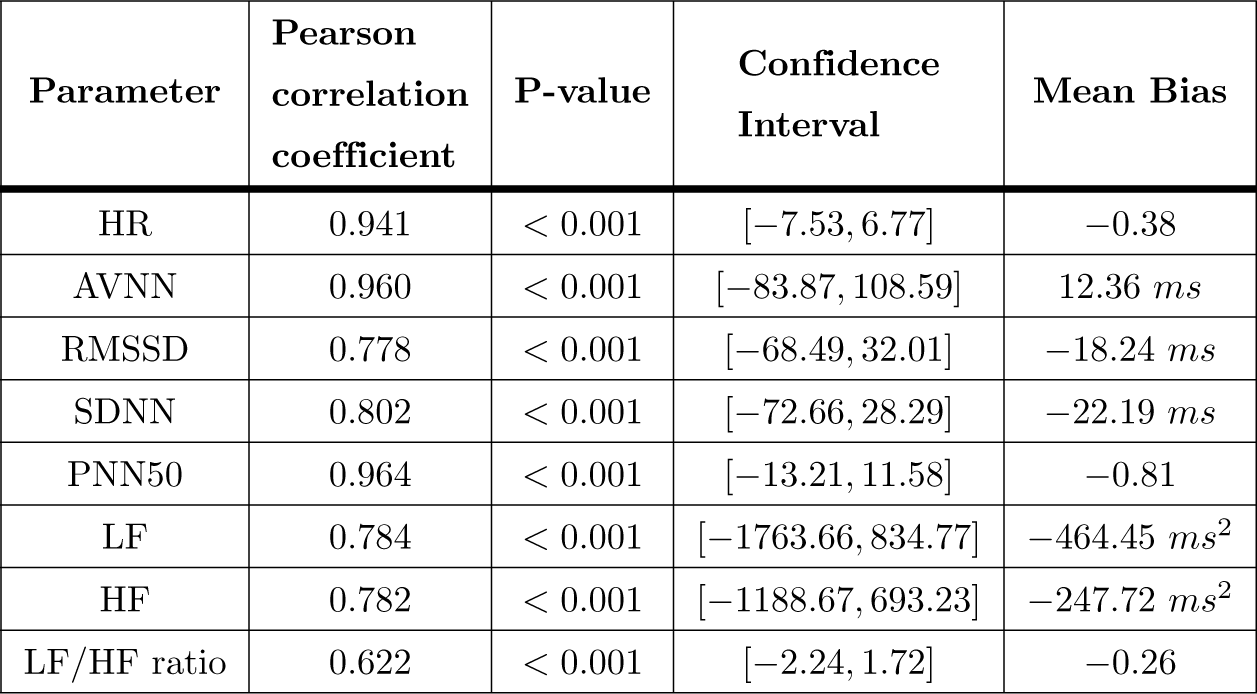
Pearson correlation coefficient, P-values, 95% Confidence Interval and Mean difference between smartwatch and Shimmer3 HR and HRV parameters in 5-minute window during sleep

The regression analysis was used to compare the accuracy of the extracted parameters from the Samsung smartwatch against the reference ECG. Fig 3 illustrates the HR and HRV parameters collected by the Samsung smartwatch (PPG) and Shimmer3 (ECG). The regression analysis was performed for the five-minute segments, and the regression lines (in red) are indicated. There are also *y* = *x* lines (in black), representing the best outcome if the PPG and ECG values are equal. The *r*^2^ values are given, indicating the scatter of the data around the regression lines. As shown in Fig 3, the fitted lines of the HR, AVNN, and pNN50 closely follow ideal lines, and their *r*^2^ values are considerably high. However, the regression lines of RMSSD, SDNN, LF, and HF relatively diverge, and their corresponding *r*^2^ values are moderate. In contrast with the other parameters, *LF/HF* data points are dispersed, and its *r*^2^ value is lower than the others.

**Fig 3.**
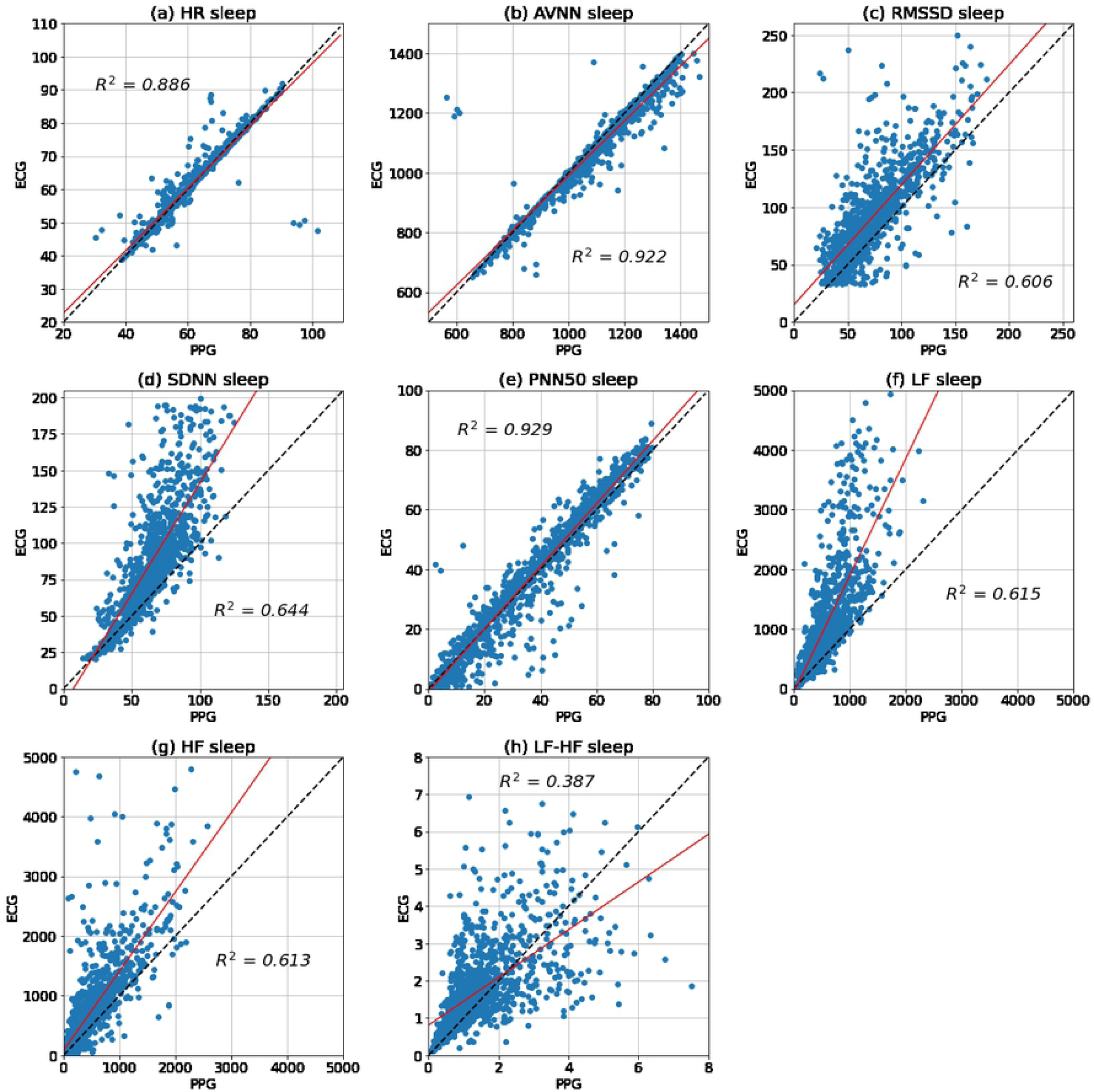
The scatter plots and regression analysis of the HR, AVNN, RMSSD, SDNN, PNN50, LF, HF, and LF/HF collected from the Samsung smartwatch and Shimmer ECG in 5-minute segments during the sleep time. The regression lines and ideal lines are indicated in red and black colors, respectively.

In addition, the Bland-Altman analysis was carried out to determine the agreement of the parameters extracted from the Samsung smartwatch and the reference ECG. The 95% confidence intervals and the mean biases are given in Fig 4 and Table 3. The results show that the smartwatch underestimates AVNN values (on average) but overestimates other parameters. In addition, there is a narrow 95% confidence interval for HR, RMSSD, SDNN, and PNN50; however, AVNN, LF, HF, and *LF/HF* ratio have relatively wider confidence intervals.

**Fig 4.**
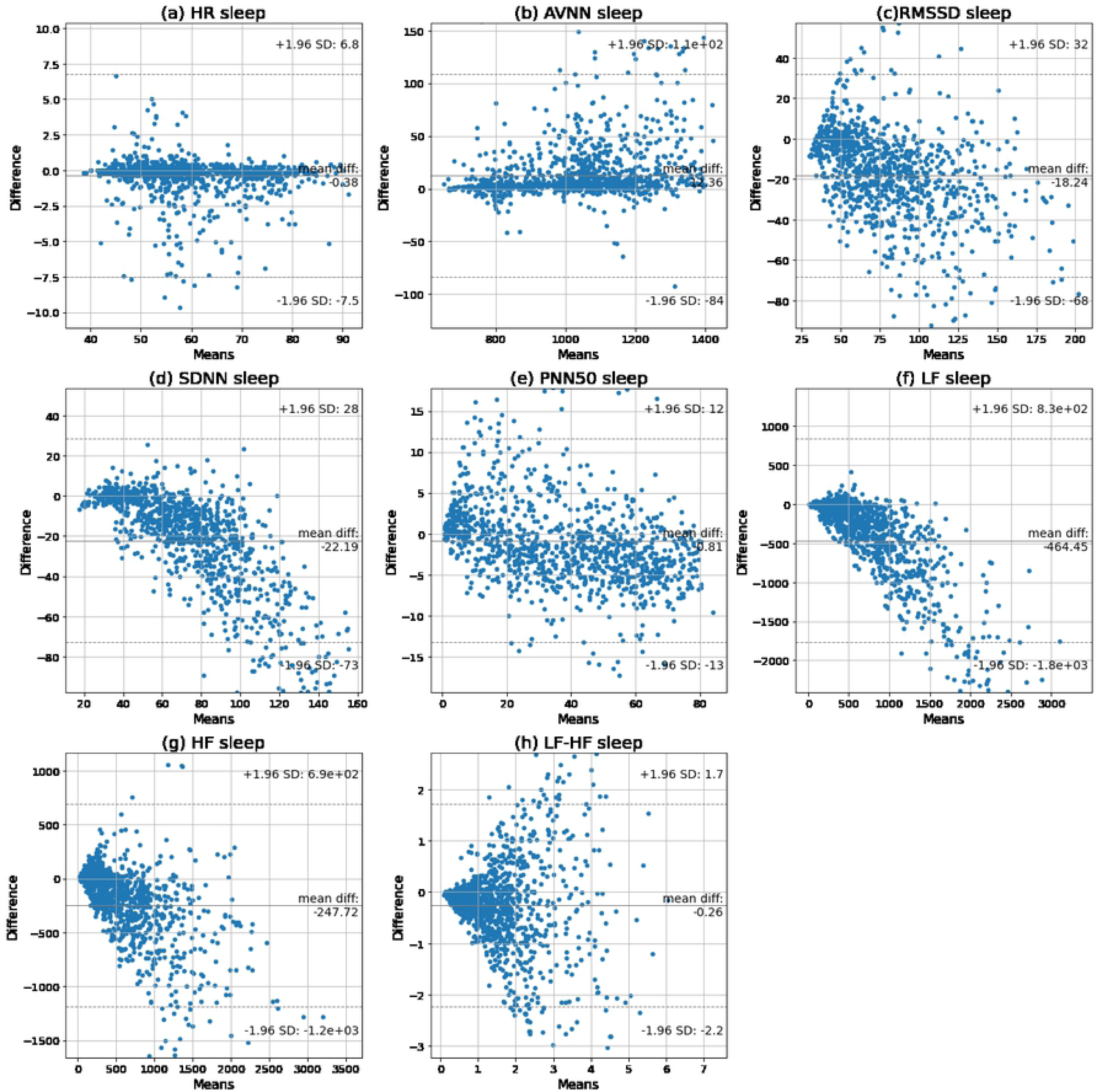
Bland-Altman plots of the HR, AVNN, RMSSD, SDNN, PNN50, LF, HF, and LF/HF in 5-minute segments obtained by smartwatch and Shimmer3 during sleep.

### Comparisons of HR and HRV parameters of the Samsung smartwatch and Shimmer3 in 5-minute time windows during awake time

This section describes the comparison of the Samsung smartwatch and the reference ECG during awake time. The awake time was obtained by excluding the sleep time from 24-hours. The assessment was performed by comparing the HR and HRV parameters in 5-minute segments. Table 4 represents the Pearson correlation coefficients along with the corresponding P-values, 95% confidence interval, and mean biases of HR and HRV parameters collected during awake time. As shown in Table 4, the results show a high positive correlation between AVNN values, a moderate positive correlation between HR values, and low positive correlations of the other HRV parameters (i.e., RMSSD, SDNN, LF, HF, and LF/HF ratio) during awake time.

**Table 4.** The calculated Pearson correlation coefficient, P-values, 95% Confidence Interval, and Mean difference between the smartwatch and Shimmer3 HR and HRV parameters in 5-minute window slots in awake time.

| Parameter | Pearson correlation coefficient | P-value | Confidence Interval | Mean Bias |
| --- | --- | --- | --- | --- |
| HR | 0.675 | <0.001 | [-43.58 , 24.65] | -9.47 |
| AVNN | 0.833 | <0.001 | [-135.12 , 254.44] | 59.66 *ms* |
| RMSSD | 0.251 | <0.001 | [-79.59 , 91.89] | 6.15 *ms* |
| SDNN | 0.404 | <0.001 | [-76.99 , 61.36] | -7.81 *ms* |
| PNN50 | 0.277 | <0.001 | [-26.0 , 60.99] | 17.5 |
| LF | 0.350 | <0.001 | [-1727.72 , 1402.18] | -162.77 *ms*<sup>2</sup> |
| HF | 0.130 | <0.001 | [-1215.82 , 1599.31] | 191.75 *ms*<sup>2</sup> |
| LF/HF ratio | 0.211 | <0.001 | [-3.38 , 2.12] | -0.63 |

We used regression analysis to compare the accuracy of the Samsung smartwatch with the ECG device. Fig 5 illustrates the regression line (in red) of the HR and HRV parameters of the five-minute segments. In addition, the *y* = *x* line is shown in these plots, which indicate the highest accuracy when the watch’s parameters are equal to the golden standard values. Fig5 also shows the *r*^2^ values, which show the scatter of data around the regression lines. The fitted lines of AVNN and HR are close to the ideal line, and their corresponding *r*^2^ values are high. However, the data points of the other HRV parameters, including RMSSD, SDNN, pnn50, LF, HF, and LF/HF ratio, are dispersed, and their *r*^2^ values are low.

**Fig 5.**
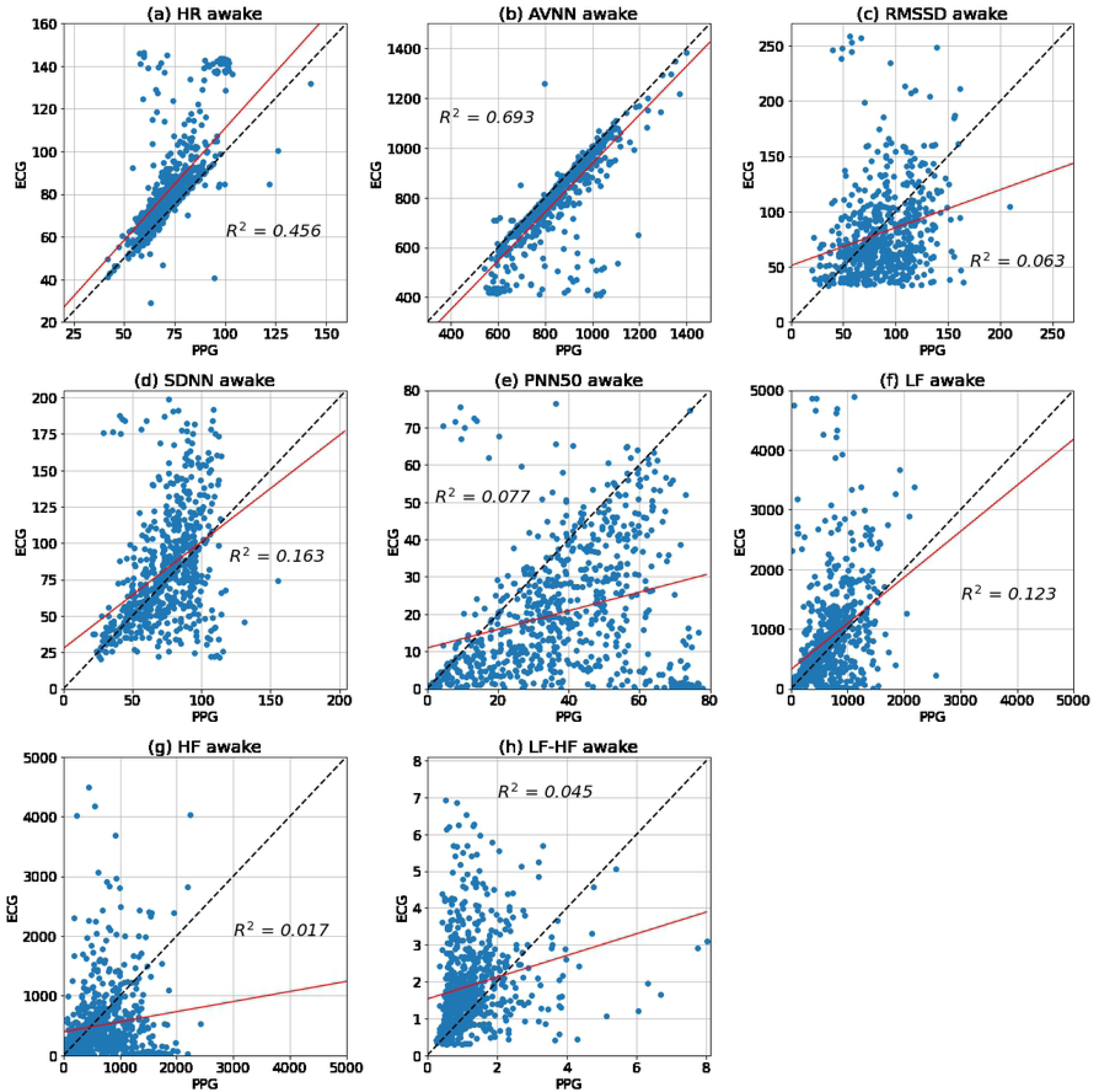
The scatter plots and regression analysis of the HR, AVNN, RMSSD, SDNN, PNN50, LF, HF, and LF/HF ratio collected from the Samsung smartwatch and Shimmer ECG in 5-minute segments during awake time. The regression and ideal lines are indicated in red and black, respectively.

We also utilized the Bland-Altman analysis to examine the agreement between the HR and HRV values during awake time. The mean biases and 95% confidence intervals are indicated in Fig 6 and Table 4. The results show that, on average, the Samsung smartwatch overestimates AVNN, RMSSD, PNN50, and HF, while it underestimates HR, SDNN, LF, and LF/HF ratio during awake time. Moreover, the 95% confidence intervals of the HR and HRV parameters are relatively wide.

**Fig 6.**
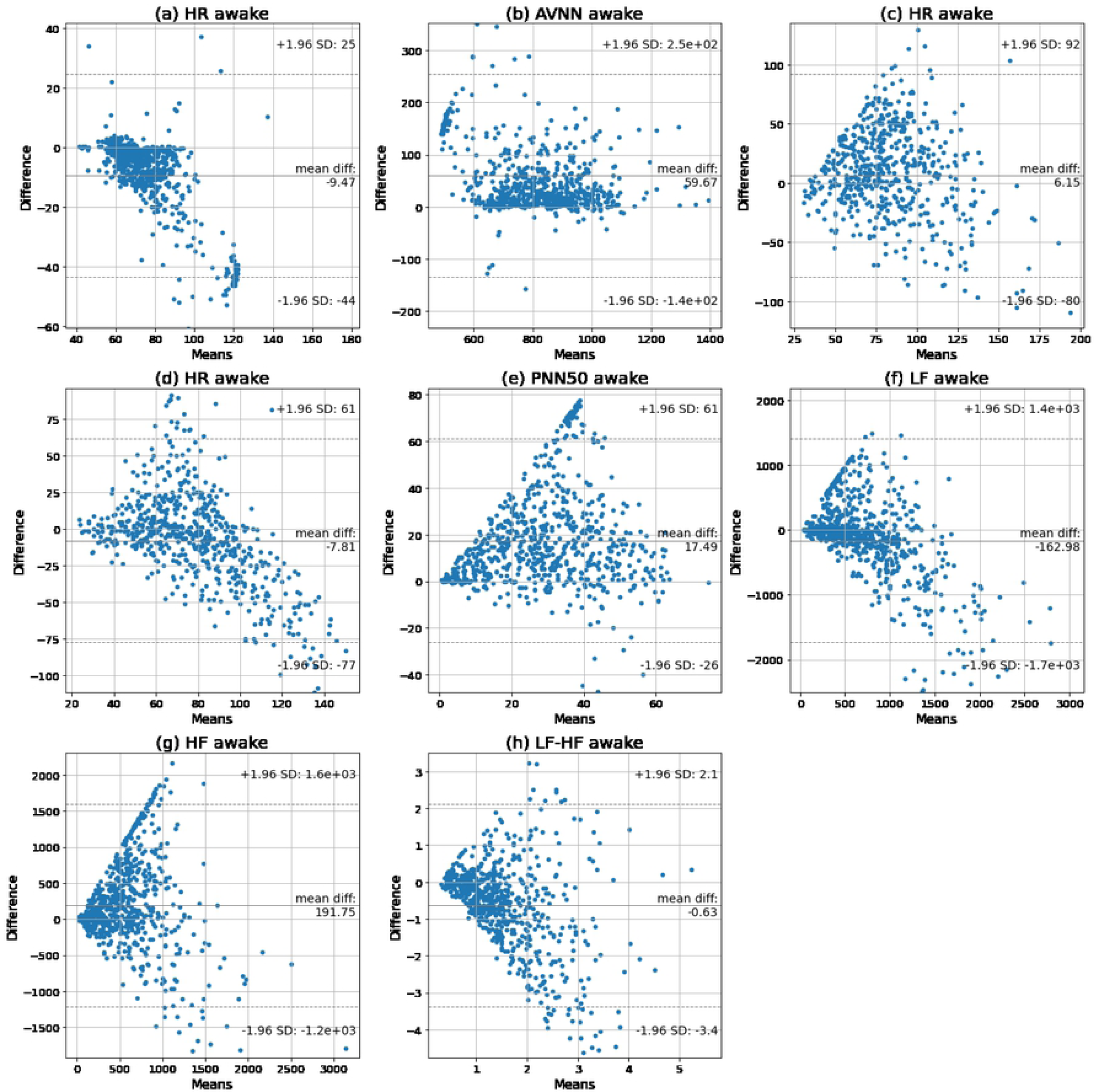
Bland-Altman plots of the HR, AVNN, RMSSD, SDNN, PNN50, LF, HF, and LF/HF in 5-minute segments obtained by the smartwatch and Shimmer3 during awake time.

## Discussion

### Principle results

In this paper, we validated the accuracy of HR and HRV parameters extracted from PPG signals collected by the Samsung smartwatch during sleep and awake time. We used short-term HRV analysis, in which HRV parameters are obtained from five-minute PPG signals [22]. Our findings during sleep time show very low mean biases of HR and AVNN, relatively low mean biases of RMSSD, SDNN, pNN50, and LF/HF ratio, and moderate mean biases of LF and HF. During the awake time, the mean biases of RMSSD and SDNN are relatively low, while the biases of HR and other HRV parameters are moderate.

Moreover, HR, AVNN, RMSSD, SDNN, pNN50, LF, and HF extracted from the Samsung watch indicated high positive correlations, while LF/HF ratio showed a moderate positive correlation with the baseline during sleep time. However, during awake time, AVNN has a high positive correlation, HR has a moderate positive correlation, and the other HRV parameters have low positive correlations with the baseline. The Samsung smartwatch underestimates HR, SDNN, LF, and LF/HF ratio but overestimates AVNN during sleep time and awake time. Moreover, the watch underestimates RMSSD, pNN50, and HF during sleep time, although it overestimates these parameters during awake time.

The error variance of the parameters is higher during awake time compared with sleep time. During sleep time, the HR, AVNN, and pNN50 have relatively low error rates, RMSSD, SDNN, LF, and HF have moderate error rates, and LF/HF ratio has a high error rate. However, during awake time, AVNN has a moderate error rate, and HR and other HRV parameters have high error rates.

In conclusion, our findings show high accuracy of HR, AVNN, and pNN50 during sleep. RMSSD, SDNN, LF, and HF have satisfactory accuracy during sleep. However, during awake time, only AVNN and HR have acceptable accuracy. HRV collection – using the Samsung smartwatch during daily activities – requires 1) noise cancellation techniques [38] to improve the signal quality and/or 2) signal quality assessment techniques [39, 40] to ensure the collected signal is not distorted and subsequently the parameters’ accuracy is acceptable.

### Comparison with previous studies

To the best of our knowledge, this is the first paper validating the HR and HRV parameters extracted from PPG signals of a smartwatch in free-living conditions. Previous studies showed high accuracy and low bias in HR extracted from wristbands including Empatica E4, Apple watch, Microsoft band, Fitbit, Garmin, PulseOn, Basis Peak, and the Wavelet wristband compared to ECG results [16–21, 24, 25, 27]. Our results also show high correlation and low bias in HR results during sleep. However, in comparison with the previous works, our results show a lower correlation of HR during the awake time when participants are involved in different activities.

Other studies validated HRV parameters during rest and specific activities. They showed the high accuracy of HRV parameters extracted from the Empatica E4 wristband for different population groups during rest and non-movement conditions [20, 24, 25]. The authors in [27] showed high correlations in SDNN and RMSSD extracted from the Wavelet wristband compared with the golden standard while resting in seated positions. Our results show a slightly lower correlation during sleep compared with previous results.

Moreover, previous studies indicated poor agreement of HRV parameters during activities for Empatica E4 [26, 41]. These studies also showed that the reliability of HRV parameters decreases with an increase in the intensity of activity. Their results follow our findings, showing higher accuracy during sleep time and lower accuracy during awake time.

Previous studies also indicated that time-domain HRV parameters have higher accuracy compared with the frequency-domain parameters. For example, the results in [23] showed high agreements of time-domain parameters during rest and mental stress but lower agreements of frequency-domain parameters for the apple watch. Microsoft Band 2 also had a higher error rate in LF/HF compared to time-domain parameters during rest and activity [26]. The results are in accordance with our results showing higher accuracy in time-domain parameters compared with frequency-domain parameters during sleep and awake time. In addition, the results in [20, 23] indicated that LF has higher accuracy than HF during the activity, which is in accordance with our results.

### Limitations

This study is limited to 24-hours data collection in everyday life settings. In the future, we will consider validating the HR and HRV parameters extracted from the smartwatch in a longer data collection period (e.g., several days or weeks). Therefore, the assessment will provide a higher confidence level on the validity results of HR and HRV parameters.

Moreover, the generalizability of the results is limited to healthy populations as we only included healthy individuals in this study. Previous works showed that the accuracy of wearable devices can vary for different population group [42, 43]. Cardiovascular disorders, such as atrial fibrillation, may cause irregular heartbeats in PPG, which will affect the HRV parameters [1]. Our future work will consider validating PPG-based HRV parameters for different age groups and various health conditions.

## Conclusion

In this paper, we comprehensively assessed the validity of HR and HRV parameters extracted from PPG signals collected for 24-hours by the Samsung Gear Sport smartwatch. The data from 28 participants were included in the study. The smartwatch was compared with an ECG device placed on the user’s chest. Our results showed low mean biases of HR, time-domain HRV, and LF/HF while moderate mean biases of LF and HF during sleep. The findings also indicated low error variances of HR, AVNN, and pNN50, moderate error variances of RMSSD, SDNN, LF, and HF, and a high error variance of LF/HF ratio during sleep. Moreover, there were high positive correlations for HR, time-domain HRV parameters, LF and HF, and a moderate positive correlation of LF/HF compared with the baseline parameters during sleep.

During the awake time, RMSSD and SDNN had low mean biases, while the other parameters showed moderate mean biases. Our findings indicated a low error variance of AVNN and a moderate error variance of HR, while the other parameters had high error variances. In addition, AVNN had a high positive correlation with the baseline, and HR had a moderate positive correlation. However, the other parameters had low positive correlations with the baseline parameters.

The smartwatch can accurately measure HR, AVNN, and pNN50 during sleep and AVNN during awake time. Moreover, the smartwatch can provide acceptable RMSSD, SDNN, LF, and HF during sleep and HR during awake time. Future work should include the assessment of the Smartwatch’s HR and HRV parameters of various population groups with different health conditions.

## Data Availability

The informed consent signed by the participants does not allow the data to be made publicly available due to ethical restriction. The data can be shared by signing a data use agreement. The data can be obtained from the corresponding author upon request.

## Acknowledgments

The authors would like to thank Elisa Lankinen, Mohsen Saei Dehghan, Bushra Zafar, and Henrika Merenlehto for contributing to the data collection. This work was supported in part by the Academy of Finland through the SLIM Project under Grant 316810 and Grant 316811, and in part by the U.S. National Science Foundation (NSF) through the UNITE Project under Grant SCC CNS-1831918 and the D-CCC Project under grant number FW-HTF CNS-2026614.

